# Guided Relaxation-Based Virtual Reality Transiently Reduces Acute Postoperative Pain and Anxiety in a Pediatric Population

**DOI:** 10.1101/2020.09.18.20192740

**Authors:** Vanessa A. Olbrecht, Keith T. O’Conor, Sara E. Williams, Chloe O. Boehmer, Gilbert W. Marchant, Susan M. Glynn, Kristie J. Geisler, Lili Ding, Gang Yang, Christopher D. King

## Abstract

**Background:** Virtual reality (VR)-based immersive games and content can distract or redirect attention. Distraction-based therapies, such as VR, have been used to reduce pain from acutely painful procedures. However, it is unlikely that distraction alone can produce the prolonged pain reduction required to manage sustained postoperative pain. Integration of VR with other pain reducing therapies, including mind-body techniques, may enhance their clinical impact. Slow breathing and relaxation techniques are used clinically to reduce pain in children. Incorporating techniques such as these into the immersive audio-visual VR experience has the potential to produce synergistic effects. The current pilot study assessed the ability of a single guided relaxation-based VR (VR-GR) session to decrease acute postoperative pain and anxiety in children and adolescents. We also explored whether pain catastrophizing and anxiety sensitivity influenced the ability of VR-GR to reduce these outcomes.

**Methods:** A total of 51 children and adolescents (ages 7-21 years) with postoperative pain followed by the Acute Pain Service at Cincinnati Children’s Hospital Medical Center were recruited over an 8-month period to undergo a single VR-GR session. Prior to VR, patients completed pain catastrophizing (PCS-C) and anxiety sensitivity (CASI) questionnaires. The primary outcome was changes in pain intensity following VR-GR (immediately, 15, and 30 minutes). Secondary outcomes included changes in pain unpleasantness and anxiety.

**Results:** Based on mixed effects models, VR-GR decreased pain intensity immediately (*p* < 0.001) and 30 minutes (*p* = 0.04) after the VR session, but not at 15 minutes (*p* = 0.16) post-session. Reductions in pain unpleasantness were observed during all time intervals (*p* < 0.001 at all intervals). Anxiety was reduced immediately (*p* = 0.02) but not at 15- (*p* = 0.08) or 30- (*p* = 0.30) minutes following VR-GR. Adjustment for covariates showed that patients with higher CASI reported greater reductions in pain intensity (*p* = 0.04) and unpleasantness (*p* = 0.01) following VR-GR. Pain catastrophizing did not impact changes in pain and anxiety following the VR session (all *p*’s > 0.10).

**Conclusion:** A single, short VR-GR session produced immediate and acute reductions in pain intensity, pain unpleasantness, and anxiety in children and adolescents with acute postoperative pain. These results encourage future randomized clinical trials to compare the effectiveness of VR-GR and mind-body based treatments to reduce postoperative pain outcomes and to reduce requirements for opioid medications during this period.

**Key Points Summary:** *Question:* Can guided relaxation-based VR transiently reduce pain and anxiety in children and adolescents following surgery?

*Findings:* A single session of guided relaxation-based VR transiently reduces pain intensity, pain unpleasantness, and anxiety in children and adolescents with severe, acute postoperative pain.

*Meaning:* Guided relaxation-based VR offers an innovative, nonpharmacologic strategy to help manage pain and anxiety in children and adolescents after surgery and combining traditional mind-body therapies with the immersive nature of VR opens new possibilities for multimodal analgesia.

## 1. Introduction

Ineffective postoperative pain management has severe long-term consequences, including increased morbidity, poorer physical functioning, longer recovery, and increased costs.^1,2^ Despite the widespread implementation of multimodal analgesia, pediatric postoperative pain remains difficult to adequately manage,^3,4^ increasing the risk of persistent postoperative pain.^5,6^ Existing studies of pediatric postoperative patients identified an approximate 20% incidence of persistent pain beyond that expected from surgery.^7^ While 80% of these patients recovered within one month, 20% maintained a reduced quality of life.^7^

Children and adolescents are at particular risk of long-term opioid abuse^8^ - as few as five days of opioid use increases this risk.^9^ While current multimodal analgesia protocols focus on regional analgesia and non-opioid medications, opioids remain ubiquitous in pain management. A recent retrospective study of opioid-naïve pediatric surgical patients found persistent opioid use in 4.8% of adolescents versus 0.1% in a matched, non-surgical cohort.^5^ Other studies found more than 25% of patients transitioned to chronic use following postoperative opioid consumption.^10^

There remains a critical need for novel, nonpharmacologic pain management strategies. Virtual reality (VR) is an innovative, nonpharmacologic pain management adjunct. VR provides an immersive, multisensory, three-dimensional environment that enables individuals to have modified experiences of reality by creating a sense of “presence.” This “sense of presence” makes VR an excellent form of distraction-based therapy.^11^ VR has been used for acute procedural, postoperative, and labor pain management. However, the majority of these studies use VR to reduce pain by redirecting attention (e.g. distraction).^12-23^ While these transient reductions are sufficient for short-term reductions in pain, they are not sufficient to treat prolonged acute pain experiences,^24,25^ including postoperative pain.

Nonpharmacologic alternatives that utilize mind-body based therapies delivered in a traditional format, like relaxation and slow breathing, have been shown to decrease anxiety and pain in children undergoing surgery.^26^ Combining traditional mind-body therapies, such as relaxation and slow breathing with immersive VR, opens new possibilities for multimodal analgesia to maximize pain control while minimizing opioid consumption.

This pilot study assessed the ability of guided relaxation-based VR (VR-GR) to decrease acute postoperative pain intensity (*p*rimary outcome) and to assess the influence of trait pain catastrophizing and anxiety sensitivity on VR-GR’s ability to reduce pain. We further assessed the ability of VR-GR to reduce pain unpleasantness and anxiety (secondary outcomes). We hypothesized that one VR-GR session would transiently reduce pain intensity, pain unpleasantness, and anxiety, with greatest reductions in patients with high baseline pain catastrophizing and anxiety sensitivity.

## 2. Methods

This single-center, prospective, pilot, clinical study in a broad pediatric postoperative population experiencing significant pain was designed to determine the ability of a single VR-GR session to transiently decrease acute postoperative pain and anxiety and assess the impact of pain catastrophizing and anxiety sensitivity on VR-GR’s pain reduction efficacy. Figure 1 shows the study design.

**Figure 1.**
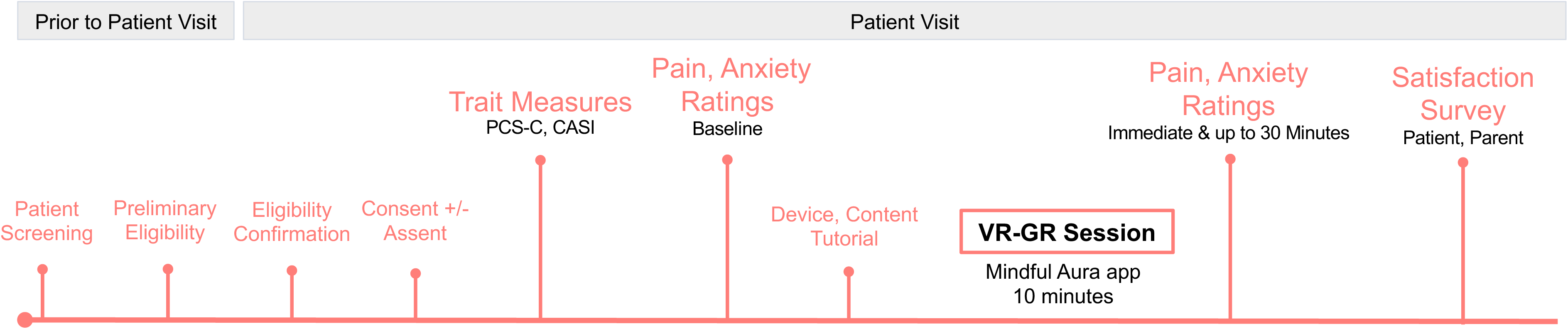
Diagram of current study.

### 2.1 Patients

From July 2019 through March 2020, we prospectively recruited 51 children and adolescents who underwent surgery and were followed postoperatively by the Acute Pain Service at Cincinnati Children’s Hospital Medical Center (CCHMC), a tertiary care, academic, pediatric hospital. This study was approved by the CCHMC Institutional Review Board (IRB #2018-2892) and was conducted in accordance with rules and regulations for ethical research. Written consent (and assent for patients ≥ 11 years) was obtained from the parent/legal guardian of patients under 18 years of age and from adolescents 18 and older. This manuscript adheres to the applicable CONSORT guidelines.

#### 2.1.1 Inclusion Criteria

Patients were eligible if they were between the ages of 7 and 21 years; if they were able to read, speak, and write English; and if they had major surgery resulting in significant postoperative pain necessitating care by the Acute Pain Service.

#### 2.1.2 Exclusion Criteria

Patients were excluded if they did not fall within the age criteria; if they did not read, speak, or write English; if they had a history of developmental delay, neurological conditions (seizure disorder, vertigo, dizziness, or significant motion sickness/nausea/vomiting), or uncontrolled psychiatric conditions; or if they had surgery of the head or neck that precluded VR use.

#### 2.1.3 Patient Information

Prior to VR-GR, we collected patient age, sex, race, surgery type, and American Society of Anesthesiologists (ASA) status.

### 2.2 Measures

The primary outcome was change in pain intensity after VR-GR. Secondary outcome measures included changes in pain unpleasantness and anxiety. See below.

#### 2.2.1 Pain Intensity, Pain Unpleasantness, and Anxiety

Pain intensity, pain unpleasantness, and anxiety (Cronbach’s α = 0.92, 0.93,0.95, respectively) were measured before and after VR-GR using the Numerical Rating Scale (NRS).^27^ A standardized script was used to explain the difference between pain intensity and pain unpleasantness. Pain was described as analogous to listening to music on the radio. Pain intensity was described as the volume of the music playing. Pain unpleasantness was related to how much the individual disliked what was playing.^28^ For each measure, patients were asked to rate symptom severity from 0 to 10 (0 = non-existent, 10 = the most severe pain/anxiety).

#### 2.2.2 Pain Catastrophizing

The Pain Catastrophizing Scale for Children (PCS-C), a validated, self-reported questionnaire assessing pain catastrophizing tendencies,^29^ was completed before the VR-GR session to determine if trait levels of catastrophizing impacted pain intensity and unpleasantness. Patients used a 5-point Likert scale to rate 13 items assessing rumination, magnification, and helplessness related to thoughts about pain. Summary scores are interpreted as low= 0-14, moderate=15-25, and high ≥26.^30^ Internal reliability in the current sample was high (Cronbach’s α = 0.92).

#### 2.2.3 Anxiety Sensitivity

The Child Anxiety Sensitivity Index (CASI) questionnaire was completed prior to VR-GR to determine if trait levels of anxiety sensitivity impacted pain and pain reductions. CASI has been used in VR studies in adolescents.^18^ This validated 18-item self-report survey measures how patients perceive symptoms of anxiety. Total scores range from 18-54.^31^ Internal reliability in the current sample was good (Cronbach’s α = 0.86).

#### 2.2.4 Patient Experience

Patient experience was measured with a 14-item questionnaire. Participants ranked how much they agreed with statements on a 4-point Likert scale (1 = Strongly agree, 2 = Agree 3 = Disagree, 4 = Strongly disagree). Statements included: “ *VR made it easier for me to tolerate my pain;” “I would recommend friends or family to try VR;” “I felt calmer and less anxious after having used VR;” “I felt like I was visiting the places in the displayed environment.”* Parent(s) of participants were asked to fill-out a similar survey to understand their perspective of their child’s VR experience.

### 2.3 VR Device and Guided Relaxation Methods

All patients used the Starlight Xperience VR device (https://www.starlight.org/virtual-reality/), a commercially available headset supplied by the Starlight Children’s Foundation’s Xperience program. The Starlight Xperience VR device is a customized version of the standalone Lenovo Mirage Solo with Daydream VR headset. Integrated headphones deliver audio content for a fully immersive experience. Patients interact and navigate within the VR environment using head movements and a small handheld controller. Patients used the “Mindful Aura” guided relaxation-based application during VR-GR to learn slow breathing and relaxation. Users are “transported” to an alpine meadow in the virtual world where a 10-minute relaxation narrative teaches focused slow, paced breathing by mirroring breathing rate with a floating butterfly, and movement of Aurora lights.

### 2.4 Procedures

After determining eligibility and obtaining consent (and/or assent where appropriate), patients completed questionnaires for trait catastrophizing (PCS-C) and anxiety sensitivity (CASI). Following completion, before VR-GR, patients rated their baseline pain intensity, pain unpleasantness, and anxiety levels (NRS).

Following baseline assessments, patients were oriented to the VR headset and given a tutorial on using the device and the application. They were instructed to remove the headset if discomfort or nausea ensue. After completing the 10-minute “Mindful Aura” session, the VR device was removed. Pain intensity, pain unpleasantness, and anxiety were recorded immediately after the VR session and 15 and 30 minutes after the experience. Patients completed a 14-item experience questionnaire after completing the study session. Parents were given a similar survey.

### 2.5 Statistical Analysis

All statistical analyses were performed using SAS 9.4 (the SAS Institute, Cary, NC). A *p* value of 0.05 was the cutoff for statistical significance. Statistical significance with Bonferroni adjustment for multiple comparison for the primary outcome (change from baseline pain intensity at three time points after VR-GR) was assessed. AR (1) – first-order autoregressive covariance structure was used in all mixed effects models.

#### 2.5.1 Descriptive Analysis

Descriptive statistics were calculated for all baseline variables and change from baseline for outcome variables. Mean, standard deviation, and/or median and interquartile range (IQR) were used for continuous variables. Frequency and percentage were used for categorical variables.

#### 2.5.2 Changes in Pain and Anxiety Following VR-GR

Pain intensity, unpleasantness, and anxiety after VR-GR were compared to baseline using paired tests (t-test or signed-rank, as appropriate) at individual time points. Changes from baseline were analyzed with mixed effects models where time (immediate, 15-, or 30-minutes after VR-GR) was used as a categorical fixed effect, with and without adjustment for other covariates (age, sex, race, ASA status, and postoperative day), as well as pain catastrophizing and anxiety sensitivity.

#### 2.5.3 Associations in Baseline Outcomes

To test the association between baseline pain catastrophizing and anxiety sensitivity with baseline outcomes (*p*ain intensity, pain unpleasantness, and anxiety), Pearson or Spearman correlation coefficients were derived as appropriate.

#### 2.5.4 Impact of Psychological Factors on Changes in Pain and Anxiety

Mixed effects models were used to examine the above associations and effect of PCS-C and CASI on changes from baseline of the outcome NRS variables, where time (immediate, 15-, or 30-minutes after VR-GR) was used as a categorical fixed effect, with and without adjustment for other covariates (age, sex, race, ASA status, and postoperative day).

#### 2.5.5 Missing Data

Missing data were examined, and all available data were used in statistical analysis.

## 3. Results

### 3.1 Baseline Participant Characteristics

We enrolled 51 patients over the 8-month recruitment period. All patients (n=51) completed pain and anxiety assessments at baseline and immediately following the VR-GR session; 100% (n=51) completed the pain assessment at 15 minutes following VR-GR and 98% (n=50) completed the anxiety assessment at 15 minutes; 88% (n=45) completed the assessments at 30 minutes following VR-GR. Missing data resulted from limitations in the clinical environment, including patients leaving for imaging studies, being seen by or receiving care from the care team, or falling asleep.

Table 1 displays the study population demographics and scores of baseline measures. Patients were mostly adolescent, male, and Caucasian. Of the 51 patients recruited, 19 (37.3%) underwent abdominal surgery, 21 (41.2%) underwent Nuss repair of pectus excavatum or chest surgery, and 11 (21.6%) underwent orthopedic procedures (such as posterior spinal fusion or major hip surgery). Half of the patients were ASA status I/II or II/IV. Patients reported moderate levels of pain intensity and unpleasantness and mild levels of anxiety prior to VR-GR (Table 1). Patients had moderate pain catastrophizing on the PCS-C and average anxiety sensitivity on the CASI ^30^. Higher PCS-C scores were associated with higher baseline anxiety prior to VR-GR (Spearman correlation coefficient=0.41, *p*=0.0029) (Supplementary Table 1). Pain catastrophizing and anxiety sensitivity were not associated with pain intensity or pain unpleasantness. Mean pain and anxiety ratings are presented in Figure 2.

**Table 1.** Demographic, Survey, and Medical Data from Study Participants. Abbreviations: PCS-C, Pain Catastrophizing Scale for Children; CASI, Child Anxiety Sensitivity Index; ASA, American Society of Anesthesiologists. Note: ASA status was not collected on three (n=3) patients.

| **Variable** | **Mean (SD) or**<br>**Number (%)** |
| --- | --- |
| **Age (years)** | 14.6 (3.2) |
| **Sex** |  |
| *Male* | 32 (63%) |
| *Female* | 19 (37%) |
| **Race** |  |
| *Caucasian* | 41 (80%) |
| *Non-Caucasian* | 10 (20%) |
| **Surgery Type** |  |
| *Pectus/Chest* | 21 (41%) |
| *Abdominal* | 19 (37%) |
| *Orthopedic* | 11 (22%) |
| **ASA Physical Status\*** |  |
| *I/II (Healthy, Mild Systemic Disease)* | 24 (47%) |
| *III/IV (Severe or life-threatening Disease)* | 24 (47%) |
| **Baseline NRS Scores** |  |
| *Pain Intensity (0-10)* | 5.11 (1.74) |
| *Pain Unpleasantness (0-10)* | 5.73 (2.30) |
| *Anxiety (0-10)* | 2.05 (2.50) |
| **Psychological Factors** |  |
| *Catastrophizing (PCS-C)* | 21.6 (11.0) |
| *Anxiety Sensitivity (CASI)* | 31.2 (3.9) |

**Figure 2.**
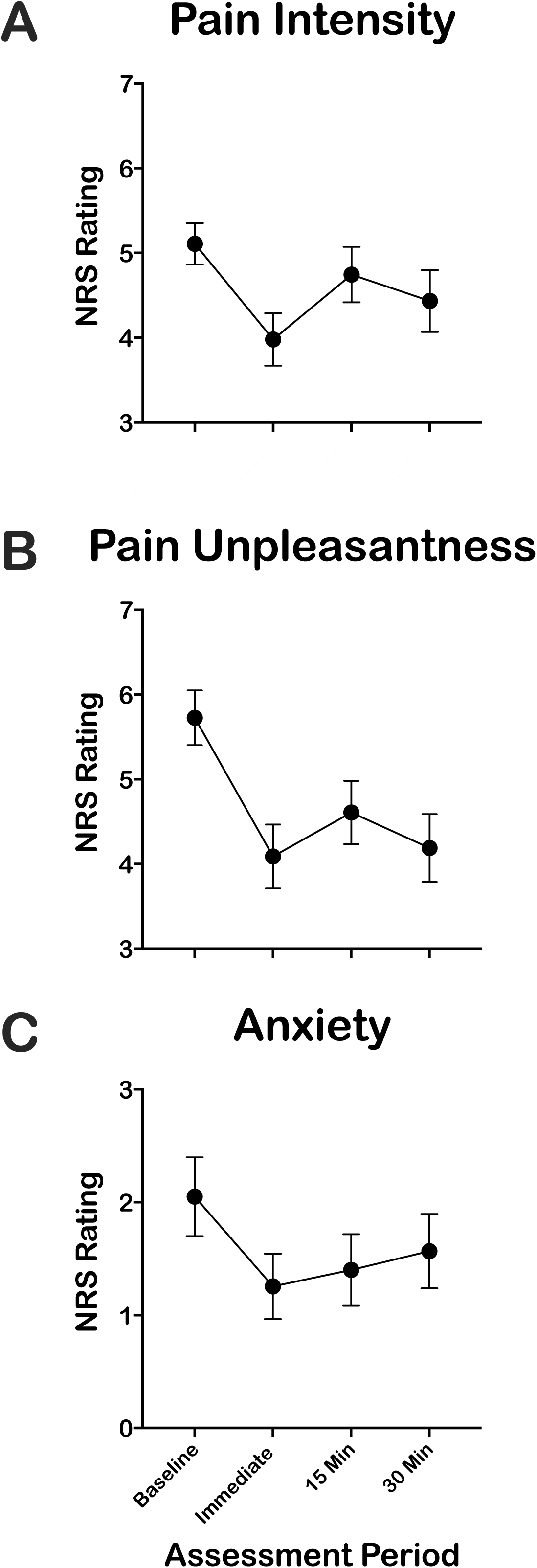
Ratings of pain and anxiety before and following VR-GR. Pain intensity (A), pain unpleasantness (B), and anxiety (C) were collected before and up to 30 minutes following VR-GR with the NRS. All patients (n=50) provided NRS scores at baseline and immediately after VR-GR. NRS scores were collected in 50 (98%) and 45 (88%) patients at 15- and 30-minutes following VR-GR, respectively. This figure is provided for graphical purposes only.

### 3.2 Primary Outcome - Pain Intensity

VR-GR had moderate to large effects on pain intensity (Figure 2A). Wilcoxon signed-rank test (Table 2) showed that pain intensity decreased immediately (Median (IQR) = -1.00 (−2.00, 0), *p*< 0.0001) following VR-GR and remained significant at 15 minutes (Median (IQR) = 0 (−1.00, 0.50), *p*=0.0275) and 30 minutes (Median (IQR) = 0 (−1.50, 0), *p*=0.0205). The mixed effects model with adjustment for covariates showed pain intensity reduction was statistically significant versus baseline when assessed immediately (LSM (SE) = -1.19 (0.25), *p* < 0.0001) and 30 minutes (LSM (SE) = -0.55 (0.26), *p*=0.0392) after VR-GR (Table 3). The decrease in pain intensity immediately after VR-GR remained significant after adjustment for multiple covariates.

**Table 2.**
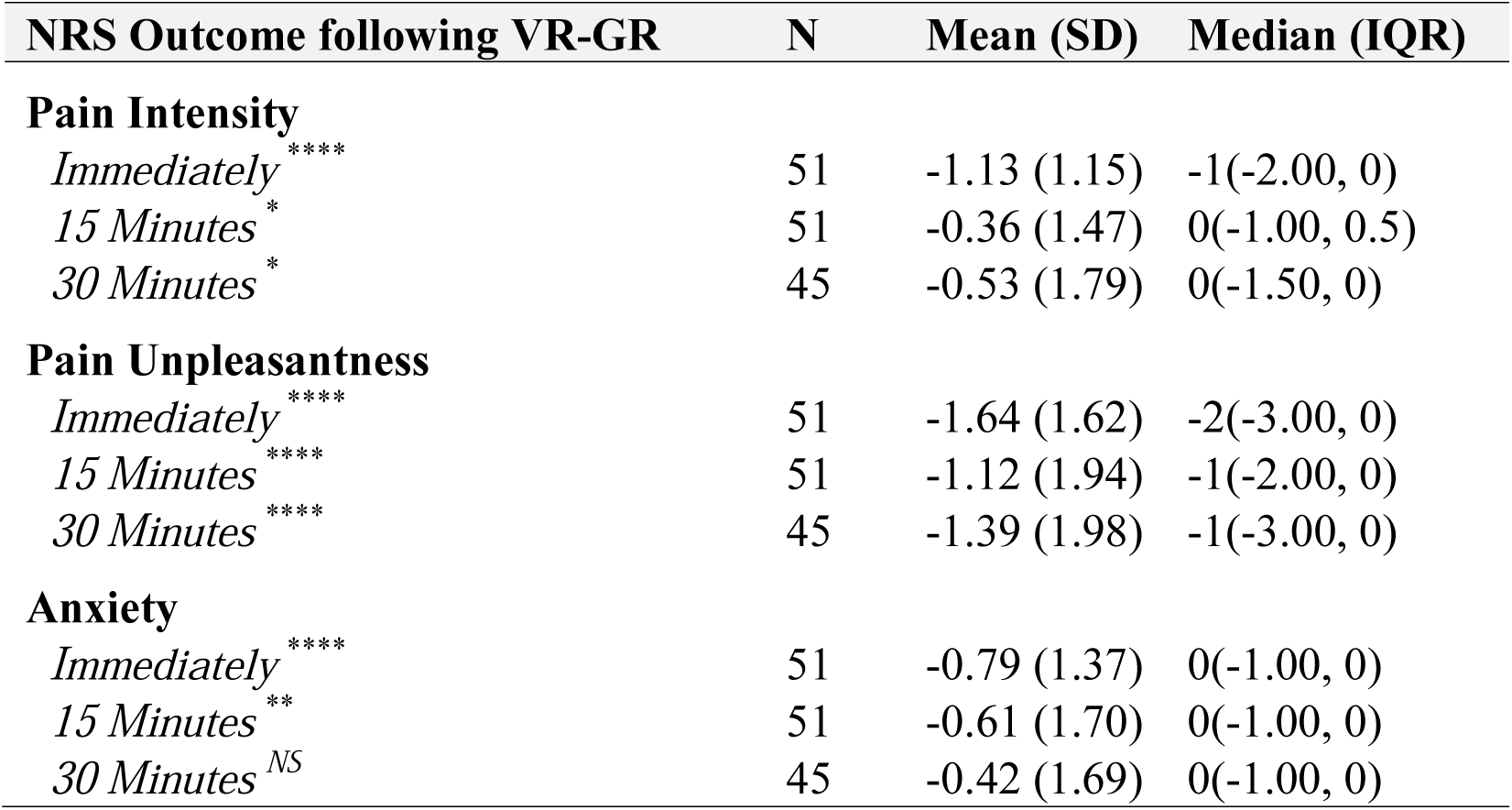
Mean (SD) and Median (IQR) Changes in Pain Intensity, Pain Unpleasantness, and Anxiety NRS Scores Following VR-GR. Wilcoxon signed-rank test was used to compared changes. Abbreviations: ^NS^, Not significant; * *p*<0.05; ** *p*<0.01; *** *p*<0.001; **** *p<*0.0001

| NRS Outcome following VR-GR | N | Mean (SD) | Median (IQR) |
| --- | --- | --- | --- |
| **Pain Intensity** |  |  |  |
| *Immediately* **** | 51 | -1.13 (1.15) | -1(-2.00, 0) |
| *15 Minutes* * | 51 | -0.36 (1.47) | 0(-1.00, 0.5) |
| *30 Minutes* * | 45 | -0.53 (1.79) | 0(-1.50, 0) |
| **Pain Unpleasantness** |  |  |  |
| *Immediately* **** | 51 | -1.64 (1.62) | -2(-3.00, 0) |
| *15 Minutes* **** | 51 | -1.12 (1.94) | -1(-2.00, 0) |
| *30 Minutes* **** | 45 | -1.39 (1.98) | -1(-3.00, 0) |
| **Anxiety** |  |  |  |
| *Immediately* **** | 51 | -0.79 (1.37) | 0(-1.00, 0) |
| *15 Minutes* ** | 51 | -0.61 (1.70) | 0(-1.00, 0) |
| *30 Minutes* <sup>NS</sup> | 45 | -0.42 (1.69) | 0(-1.00, 0) |

### 3.3 Secondary Outcome - Pain Unpleasantness

VR-GR also had a moderate to large effect on pain unpleasantness (Figure 2B). Wilcoxon signed-rank test (Table 2) revealed that pain unpleasantness decreased immediately following VR-GR (Median (IQR) = -2.00 (−3.00, 0), *p*< 0.0001); and remained significantly decreased at 15 (Median (IQR) = -1.00 (−2.00, 0), *p*< 0.0001) and 30 minutes (Median (IQR) = -1.00 (−3.00, 0), *p*< 0.0001) following VR-GR. In our adjusted mixed effects models (Table 3), reductions in pain unpleasantness were observed immediately (LSM (SE) = -2.07 (0.32), *p* < 0.0001), 15 minutes (LSM (SE) = -1.52 (0.32), *p*< 0.0001), and 30 minutes (LSM (SE) = -1.84 (0.33), *p*< 0. 0001) following VR-GR.

**Table 3.** Mixed-Effect Model Reflecting Changes in Pain Intensity, Pain Unpleasantness, and Anxiety NRS Scores Following VR-GR. Mixed Effects Models were run without (unadjusted) and with (adjusted) covariates. Notes: ^α^, Study covariates including demographic (age, sex, race), the time following surgery (*p*ostoperative day, POD), ASA status, and overall scores on the PCS-C and CASI. Abbreviations: ^NS^, Not significant; * *p*<0.05; ** *p*<0.01; *** *p*<0.001; **** *p<*0.0001

| **NRS Outcome following VR-GR** | **Unadjusted Model**<br>**LSM (SE)** | **Adjusted Model<sup>α</sup>**<br>**LSM (SE)** |
| --- | --- | --- |
| **Pain Intensity** |  |  |
| *Immediately* | -1.13 (0.21) **** | -1.19 (0.25) **** |
| *15 Minutes* | -0.36 (0.21) <sup>NS</sup> | -0.36 (0.25) <sup>NS</sup> |
| *30 Minutes* | -0.57 (0.22) * | -0.55 (0.26) * |
| **Pain Unpleasantness** |  |  |
| *Immediately* | -1.64 (0.26) **** | -2.07 (0.32) **** |
| *15 Minutes* | -1.12 (0.26) **** | -1.52 (0.32) **** |
| *30 Minutes* | -1.46 (0.27) **** | -1.84 (0.33) **** |
| **Anxiety** |  |  |
| *Immediately* | -0.79 (0.22) *** | -0.74 (0.31) * |
| *15 Minutes* | -0.63 (0.22) ** | -0.55 (0.31) <sup>NS</sup> |
| *30 Minutes* | -0.44 (0.23) <sup>NS</sup> | -0.33 (0.31) <sup>NS</sup> |

### 3.4 Secondary Outcome - Anxiety

We found moderate effects of VR-GR on anxiety (Figure 2C). Wilcoxon signed-rank test (Table 2) revealed anxiety decreased from baseline immediately following VR-GR (Median (IQR) = 0 (−1.00, 0), *p*< 0.0001). Anxiety remained lower at 15 minutes following VR-GR (Median (IQR) = 0 (−1.00, 0), *p*=0.0068) but was not significantly reduced 30 minutes after VR-GR. In our adjusted mixed effects models, anxiety reduction was observed immediately (LSM (SE) = -0.74 (0.31), *p*=0.0189) following VR-GR; reduction was not significant at 15 or 30 minutes (Table 3).

### 3.5 Effects of Psychological Factors on Changes in Scores

Using a series of adjusted mixed effects models, anxiety sensitivity levels were associated with greater reductions in pain intensity (Beta (SE) = -0.06 (0.03), *p* = 0.0426) and pain unpleasantness (Beta (SE) = -0.09 (0.04), *p*=0.0111). Pain catastrophizing was not associated with changes in pain and anxiety following VR-GR (Table 4).

**Table 4.** Impact of Psychological Factors on Changes in Pain Intensity, Pain Unpleasantness, and Anxiety NRS Scores Following VR-GR. Mixed Effects Models were run for PCS-C and CASI without (unadjusted) and with (adjusted) other covariates. Notes: ^α^, Study covariates including demographic (age, sex, race), the time following surgery (*p*ostoperative day, POD), ASA status, and controlling for the overall scores on the PCS-C or CASI. Abbreviations: ^NS^, Not significant; * *p*<0.05

| **NRS Outcome following VR-GR** | **Unadjusted Model**<br>**Beta (SE)** | **Adjusted Model<sup>α</sup>**<br>**Beta (SE)** |
| --- | --- | --- |
| **Pain Intensity** |  |  |
| *Pain Catastrophizing (PCS-C)* | -0.0001 (0.01)<sup>NS</sup> | 0.01 (0.02)<sup>NS</sup> |
| *Anxiety Sensitivity (CASI)* | -0.04 (0.02)<sup>NS</sup> | -0.06 (0.03)<sup>*</sup> |
| **Pain Unpleasantness** |  |  |
| *Pain Catastrophizing (PCS-C)* | 0.01 (0.02)<sup>NS</sup> | 0.03 (0.02)<sup>NS</sup> |
| *Anxiety Sensitivity (CASI)* | -0.05 (0.03)<sup>NS</sup> | -0.09 (0.04)<sup>*</sup> |
| **Anxiety** |  |  |
| *Pain Catastrophizing (PCS-C)* | -0.02 (0.02)<sup>NS</sup> | -0.004 (0.02)<sup>NS</sup> |
| *Anxiety Sensitivity (CASI)* | -0.06 (0.03)<sup>*</sup> | -0.04 (0.04)<sup>NS</sup> |

### 3.6 Study Covariates

When assessing the impact of other covariables on the effect of VR-GR, mixed effects models showed that with adjustment for other covariables, Caucasian patients had a smaller decrease in pain unpleasantness from baseline compared to non-Caucasian participants [Difference in LSM (95% CI) = 1.20 (0.11, 2.28), *p*=0.0312]. Without adjustment for covariates, older participants had a smaller decrease in pain intensity (Beta (SE) = 0.12 (0.05), *p*=0.0285). No other variables (ASA, postoperative day, or race) were associated with VR-GR.

### 3.7 Patient Satisfaction and Experience with Virtual Reality

Patients reported a very positive overall VR experience. When asked if they would recommend VR to friends and family, 96% of children strongly agreed (59%) or agreed (37%). Most patients (n=45, 88%) believed they felt *“calmer and less anxious after having used VR”* and that VR *“made it easier for [them] to tolerate [their] pain.”* Parents reported a positive overall experience when asked the same questions. Parents (n=44, 100%) who completed the experience questionnaire strongly agreed (82%) or agreed (18%) that they would recommend VR to friends or family, 41 (93%) believed VR made their child feel calmer and less anxious and 37 (84%) believed VR helped their child tolerate pain. No patients experienced any side effects.

## 4. Discussion

VR-GR may be beneficial in pediatric acute postoperative pain management. One 10-minute VR-GR session decreased pain intensity, pain unpleasantness, and anxiety in children and adolescents with acute postoperative pain following major surgery. Greatest effects were observed for pain unpleasantness, with lesser reductions in pain intensity and anxiety. Patients with higher anxiety sensitivity showed greater reductions in pain intensity and unpleasantness after VR-GR. Pain catastrophizing did not impact response. Qualitative assessments suggested patients had positive experiences with VR-GR.

Although pain management strategies increasingly incorporate multimodal analgesia, the percentage of patients experiencing severe postoperative pain has not changed in 20 years.^3,4^ Significant postoperative pain is associated with poorer functional outcomes and chronic pain development.^2,7^ Risk of chronic pain development is well studied in adults, but not children.^32^ The few pediatric studies identified an approximate 20% incidence of persistent postsurgical pain.^7^ About 80% of these children recovered one month postoperatively, but about 20% maintained pain-related quality of life reductions.

Children are at risk of persistent opioid use following surgery, but this is also not well studied. A recent retrospective study of 13 to 21-year-old opioid-naïve patients compared those who had undergone surgery and received an opioid prescription versus those that did not have surgery. Persistent opioid use was seen in 4.8% of subjects that underwent surgery and received opioids versus 0.1% in those that did not have surgery.^5^ A study of children and adolescents with chronic pain found more than 25% transitioned to chronic opioid use after receiving opioids for postoperative pain.^10^

Few studies utilized VR for acute postoperative pain in adults,^21,33^ and none used VR to manage pediatric acute postoperative pain. Most VR studies use distraction-based VR (VR-D) to temporarily reduce pain by redirecting attention.^11,13,14,16,17,19,34^ Without VR, distraction alone provides little proven pain management benefit,^35^ with no lasting significant impact on pain.^24,34^ The improved efficacy of VR-D to temporarily reduce pain is likely due to the immersive nature of VR.^33,36^ Although VR-D is more effective than distraction alone, VR-D use may be limited as transient pain reductions are insufficient to treat prolonged acute pain, including postoperative pain. Incorporation of VR with other pain reducing strategies, such as guided relaxation, may promote more sustained pain relief beyond the temporary impact of distraction.^25^

Mind-body therapies, like relaxation and guided imagery, decrease anxiety and help manage pain in children undergoing surgery.^26^ Unfortunately, their integration into postoperative clinical care is fraught with challenges, including access to care, high cost, and provider availability. Using VR to deliver these therapies can increase accessibility and enhance acceptability, motivation,^37^ and adherence in pediatric patients compared to methods without VR, making this therapy more engaging and relevant.^38^

This was the first study to integrate mind-body therapy with VR to help pediatric postoperative pain. We preliminarily assessed use of VR-GR to decrease acute postoperative pain and whether anxiety influenced VR-GR pain management efficacy in a broad, pediatric population with significant postoperative pain. “Mindful Aura” VR software taught patients to slow their breathing to downregulate their stress response, resulting in decreased pain.^39^ A single, short VR-GR session produced immediate, acute reductions in pain intensity, pain unpleasantness, and anxiety. Reductions in pain unpleasantness were observed during all time intervals. Anxiety was reduced immediately and 15-minutes following VR-GR. Patients with higher CASI showed greater reduction in pain intensity and pain unpleasantness following VR-GR. Patients with higher trait pain catastrophizing did not respond better to VR-GR.

This study had several limitations. Study design did not allow comparison of VR-GR versus another distraction-based pain management method or control. This study was designed only to test feasibility and obtain pilot data demonstrating the ability to use VR-GR in hospitalized pediatric acute postoperative patients and demonstrate transient reductions in pain and anxiety. VR is relatively new technology that requires instruction and experience prior to use. Few patients had previous VR experience prior to this study, and some had difficulty navigating menus which may have negatively impacted their VR experience. Future studies will incorporate targeted education and program orientation sessions to acclimate patients to the VR device prior to the study. This study used a single, short VR session during the postoperative period. We recognize that a single VR-GR session is unlikely to produce sustained effects on pain and anxiety and will pursue further clinical studies using repeated sessions. Increased session length or repeated sessions over several days may have resulted in increased and sustained benefit as patients became more familiar with the device and subsequently more immersed. The benefit of mind-body therapies requires multiple sessions and practice between sessions to develop mastery^39,40^. Future studies will incorporate these aspects. This study used a diverse patient population. While this lack of standardization is a limitation, each patient served as their own baseline. By studying a diverse population, we were able demonstrate that VR can be successfully used in many different patient types, expanding the generalizability of our findings. Finally, there is potential for bias when self-reporting pain scores due to expectation of treatment impact, which could have produced lower self-reported pain and anxiety scores following VR-GR. This will be addressed in future studies by addition of a control arm.

Data from this pilot study will inform design of a large, randomized clinical trial assessing VR-GR impact on acute postoperative pain and anxiety versus control and exploring effects of increased numbers of sessions in the immediate postoperative period. Our data showed most patients had very positive experiences with VR– most felt VR-GR helped reduce their anxiety and ability to tolerate pain. Positive patient experiences suggest patients would likely comply with VR treatment, further supporting its utility and viability.

In summary, our study demonstrates successful use of VR-GR for pediatric acute postoperative pain management. A single VR-GR session produced transient decreases in pain and anxiety. Future research is needed to investigate repetitive applications of VR-GR in the postoperative setting with the goal of providing greater, more sustained postoperative pain and anxiety reduction in children and adolescents.

## Data Availability

Data available upon request.

## Acknowledgments

Authors thank Maria Ashton MS, RPh, MBA, Medical Writer, Department of Anesthesia Cincinnati Children’s Hospital Medical Center, Cincinnati, Ohio for writing assistance, editing, and proofreading.

## Funding

This work was funded by an internal research grant, Research Innovation/Pilot Funding, as well as the Department of Anesthesiology at Cincinnati Children’s Hospital.

## Conflicts of Interest

None

## Abbreviated title

Guided relaxation-based virtual reality for postoperative pain and anxiety

## Author Contributions

VO: Conception of study idea, design of research protocol, implementation of protocol/procedures, writing of manuscript; KO: Design and implementation of protocol/procedures, writing of manuscript; SW: Conception of study idea, design of protocol, implementation of protocol/procedures, writing of manuscript; CB: Design and implementation of protocol/procedures, writing of manuscript; GM: Design and implementation of protocol/ procedures, writing of manuscript; SG: Design and implementation of protocol/procedures; KG: Design and implementation of protocol/procedures; LD: Study design, development of statistical analysis plan, statistical analysis, writing of manuscript; GY: Study design, development of statistical analysis plan, statistical analysis, writing of manuscript; CK: Conception of study idea, design of research protocol, implementation of study protocol/procedures, writing of manuscript. All authors revised and modified this manuscript and approved the final version.

**Glossary of Terms**
ASA: American Society of Anesthesiologists
CASI: Child Anxiety Sensitivity Index
CCHMC: Cincinnati Children’s Hospital Medical Center
NRS: Numerical rating scale
PCS-C: Pain Catastrophizing Scale for Children
VR: Virtual reality
VR-D: Distraction-based virtual reality
VR-GR: Guided relaxation-based virtual reality

---

**Supplementary Table 1.**
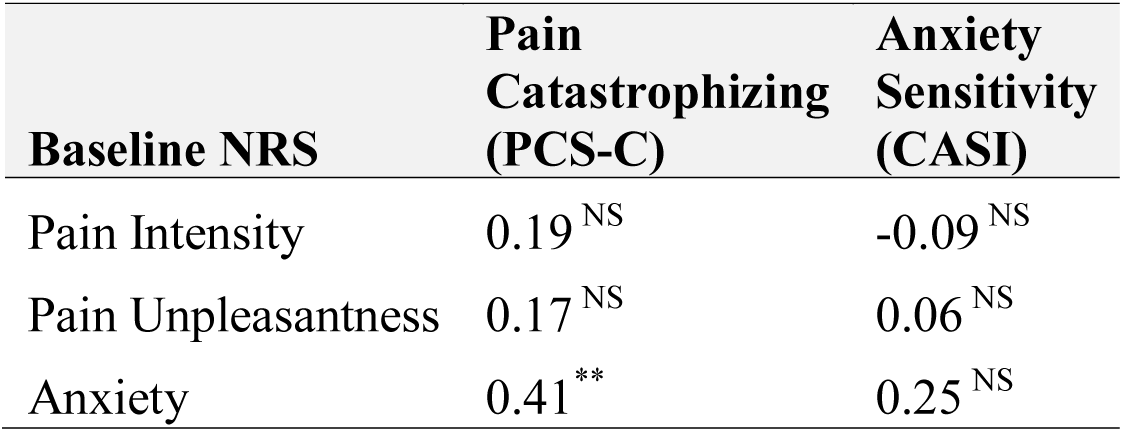
Spearman Correlation Between Baseline NRS Scores and Surveys. Abbreviations: ^NS^, Not significant; ** *p*<0.01

## Notes

### Competing Interest Statement

The authors have declared no competing interest.

### Clinical Trial

NCT04556747. This trial was registered retrospectively as it was a pilot study that involved only one group of patients without a control group and, as such, the authors were not aware it required registration.

### Funding Statement

This study was supported by the Department of Anesthesia at Cincinnati Children's Hospital Medical Center.

### Author Declarations

This study was approved by the IRB at Cincinnati Children's Hospital Medical Center.

